# Self-selected step length asymmetry is not explained by energy cost minimization in individuals with chronic stroke

**DOI:** 10.1101/19013854

**Authors:** Thu M. Nguyen, Rachel W. Jackson, Yashar Aucie, Digna de Kam, Steven H. Collins, Gelsy Torres-Oviedo

**Affiliations:** Department of Mechanical Engineering, Stanford University; Department of Bioengineering, Stanford University; Department of Bioengineering, University of Pittsburgh

**Keywords:** stroke, asymmetry, biofeedback, metabolic cost, gait

## Abstract

**Background:** Asymmetric gait post-stroke is associated with decreased mobility, yet individuals with chronic stroke often self-select an asymmetric gait despite being capable of walking more symmetrically. The purpose of this study was to test whether self-selected asymmetry could be explained by energy cost minimization. We hypothesized that short-term deviations from self-selected asymmetry would result in increased metabolic energy consumption, despite being associated with long-term rehabilitation benefits. Other studies have found no difference in metabolic rate across different levels of enforced asymmetry among individuals with chronic stroke, but used methods that left some uncertainty to be resolved.

**Methods:** In this study, ten individuals with chronic stroke walked on a treadmill at participant-specific speeds while voluntarily modulating step length asymmetry. We included only participants with significant self-selected asymmetry who were able to significantly alter asymmetry using visual biofeedback. Conditions included targeting zero asymmetry, self-selected asymmetry, and double the self-selected asymmetry. Participants were trained with the biofeedback system in one session, and data were collected in three subsequent sessions with repeated measures. Self-selected asymmetry was consistent across sessions. A similar protocol was conducted among unimpaired participants.

**Results:** Participants with chronic stroke substantially altered step length asymmetry using biofeedback, but this did not affect metabolic rate (ANOVA, p = 0.8). In unimpaired participants, self-selected step length asymmetry was close to zero and corresponded to the lowest energy cost (ANOVA, p = 6e-4). While the symmetry of unimpaired gait may be the result of energy cost minimization, self-selected step length asymmetry in individuals with chronic stroke cannot be explained by a similar least-effort drive.

**Conclusions:** Interventions that encourage changes in step length asymmetry by manipulating metabolic energy consumption may be effective because these therapies would not have to overcome a metabolic penalty for altering asymmetry.

## Background

Stroke often results in hemiparesis and gait asymmetries, such as spatial, temporal, or kinematic differences between the paretic and nonparetic legs(1–3). Asymmetric gait post-stroke has been associated with slower walking speeds(4) and higher metabolic energy consumption(5,6) compared to unimpaired walking. Conventional gait retraining by physical therapists can reduce gait asymmetries, particularly step length asymmetry, and improve speed and energy economy, but sessions are costly, limiting access. More automated rehabilitation techniques to reduce step length asymmetry have been developed using split-belt treadmills(7–9) or rehabilitation robots(10,11). These interventions, however, have not been more effective than conventional physiotherapy for individuals with chronic stroke(8,11–15). A better understanding of the mechanisms driving step length asymmetry in individuals with chronic stroke could allow for the development of more targeted, effective, and accessible gait interventions.

Unimpaired individuals self-select many gait parameters, such as step frequency(16), step width(17), and even arm swinging characteristics(18), to minimize their energy cost of walking, a strategy that might also explain self-selected asymmetries in post-stroke gait. Deviations from self-selected gait tend to lead to an increase in energy expenditure, creating bowl-like relationships, or cost landscapes, between energy cost and gait parameters, with the energy minimum at the self-selected parameter value(19). The relationship between step length asymmetry and metabolic rate has not yet been characterized in unimpaired individuals, but studies that enforce absolute differences in step length(20) or asymmetry in step time(21) suggest that they self-select nearly symmetric step lengths that correspond to a lower energy cost than asymmetric gait. Because stroke often leads to physical asymmetries, such as paretic leg muscle weakness(22), muscle spasticity(23), or reduced paretic-leg push-off force(24,25), an asymmetric gait could be metabolically optimal for individuals with chronic stroke. On the other hand, factors other than effort minimization, such as perceived effort, avoidance of fatigue, comfort, or stability, could be primarily responsible for the observed step length asymmetry in this population.

The effects of acutely changing step length asymmetry may differ from those of slowly changing step length asymmetry through the process of rehabilitation. Long-term rehabilitation interventions that decrease gait asymmetry have shown that cost of transport often improves concurrently(26). However, other effects of long-term rehabilitation, such as increased muscle strength(27) or improved motor control, may enable individuals to walk with reduced asymmetry more efficiently. Therefore, acute reductions in gait asymmetry that are not accompanied by the neuro-musculoskeletal changes often seen in long-term rehabilitation may not correlate with improvements in walking economy. Even for individuals who walk with highly asymmetric gaits, their self-selected asymmetry may be the most energy efficient one, and acute changes in gait asymmetry could still lead to increased energy consumption.

Step length asymmetry is changed acutely during split-belt walking, but this task change may also change the optimal step length asymmetry. When belt speeds are matched immediately following split-belt training, step length asymmetry is acutely changed, but this washout effect does not persist long enough to collect steady-state metabolic rate measurements(28). To determine whether self-selected gait asymmetry minimizes energy cost, gait asymmetry would need to be varied independently from other factors affecting metabolic energy consumption, within an individual participant, while metabolic rate is measured.

Previous studies measured metabolic rate while individuals with chronic stroke acutely altered step length asymmetry and step length difference using biofeedback(20,29). These studies found no difference in metabolic rate between self-selected and altered step length asymmetries in stroke survivors. However, some uncertainty remains to be resolved. Each prior study included participants with self-selected step length asymmetry values close to zero. This makes differentiating between the potential optimality of self-selected asymmetry and that of absolute symmetry difficult. Prior studies also included some participants who were unable to reliably alter step length asymmetry, so asymmetry values in different conditions could have been similar. This can reduce the power of a numerical analysis intended to identify an effect of step length asymmetry on metabolic rate. In prior studies, participants were instructed to hold onto the treadmill handrails to help with stability and minimize fall risk. However, participants could have relied more heavily on the handrails during more difficult conditions to improve stability or to help maintain the correct speed. Both improved balance(30,31) and handrail holding(32) during treadmill walking have been shown to reduce metabolic energy consumption. To minimize the amount of walking and number of experimental sessions for participants with chronic stroke, participants in these studies were familiarized with the biofeedback on the same day as the data collection. However, motor learning can have an effect on metabolic rate during a novel task; as individuals learn a new task, metabolic power(33,34) and muscle activity(35) typically decrease, with steady state reached after hours or days of practice. For example, Sánchez et al. recently showed that split-belt treadmill training takes longer than originally thought(36), and training over multiple sessions can facilitate better learning because memory consolidation occurs during sleep(37).

The purpose of this study was to characterize the relationship between step length asymmetry and metabolic energy consumption during walking in individuals with chronic stroke and unimpaired individuals. We screened for individuals with chronic stroke who exhibited clinically meaningful self-selected step length asymmetry, so as to differentiate between the potential optimality of self-selected asymmetry and perfect symmetry. Only participants who could substantially alter their asymmetry with biofeedback were included, which ensured that the effects of changes in asymmetry could be robustly analyzed. We disallowed participants from using handrails during all conditions to avoid uncertainty related to the potential benefits of improved balance or forward pulling during more difficult conditions. Participants with chronic stroke received training on the biofeedback system during the first session to facilitate task learning and ensure that all participants could alter their baseline asymmetry with biofeedback. Data were collected in three subsequent sessions. During each collection session, conditions were presented in a different order to avoid ordering effects, and the first condition of the session was repeated to reduce within-session training effects. We hypothesized that individuals with chronic stroke would self-select the step length asymmetry that minimized their metabolic energy consumption during walking, and that more symmetric or asymmetric gaits would result in a higher metabolic cost. We hypothesized that unimpaired individuals would self-select the step length asymmetry, near symmetric, that minimized their metabolic cost, and more asymmetric gaits would increase metabolic cost. The results from this study were expected to improve our understanding of the mechanisms driving self-selected step length asymmetry and influence the development of new gait retraining techniques.

## Methods

We conducted an experiment in which we asked individuals with chronic stroke and unimpaired participants to alter their step length asymmetry using biofeedback while walking on an instrumented treadmill at a participant specific speed. Participants were first familiarized with the biofeedback system and practiced walking in conditions targeting a variety of asymmetry levels. Data collection trials followed familiarization. Participants walked in each biofeedback condition for six minutes, and metabolic rate measurements were collected. Rest breaks were given between each walking bout. Participants wore a safety harness that did not provide body weight support. Participants were instructed not to hold onto the treadmill handrail unless they felt like they might lose their balance. All participants provided written informed consent before participating. The experimental protocol was approved by the University of Pittsburgh Institutional Review Board.

### Biofeedback

Biofeedback was used to enforce various step length asymmetries (Figure 1A). Participants were provided with a visual representation of the step length targets (Figure 1B). Bars on the screen grew in proportion to the forward position of the lateral malleolus of the swing leg with respect to the ankle of the standing leg. Participants were asked to heel strike onto the treadmill when the bar reached the target. Each target allowed for 2.5 cm of error either ahead of or behind the targeted step length value. As an incentive, the step length target would explode when the participant hit it (Figure 1C). If the participant missed the target, the target would turn red (Figure 1D). In both cases, a yellow line would appear to show the participant’s step length as additional feedback for the next step.

**Figure 1.**
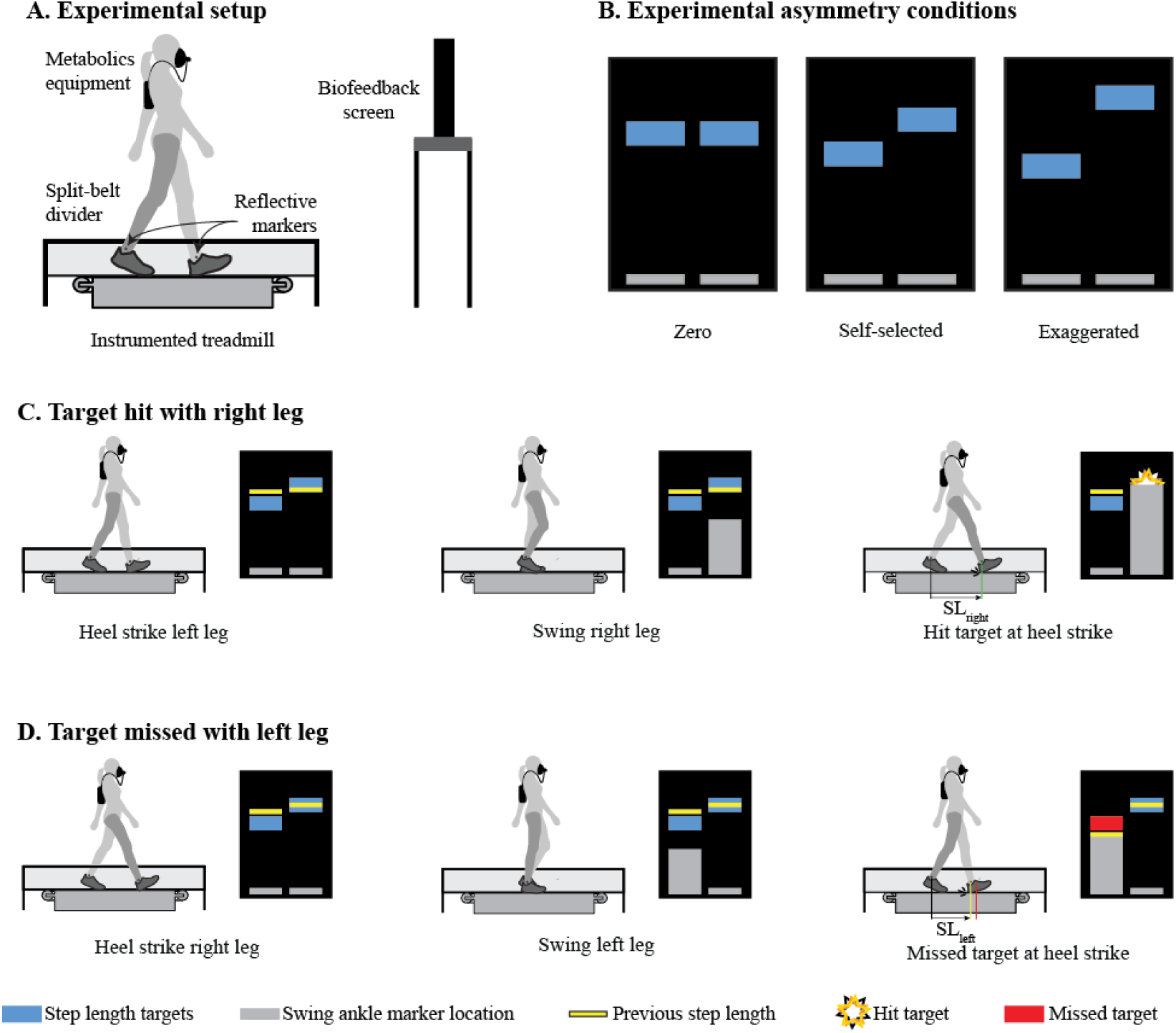
Biofeedback system. (A) Participants walked on an instrumented treadmill with a thin divider to more accurately detect left and right heel strikes. Reflective markers were attached to the ankles to measure step length at heel strike. (B) Example asymmetry conditions for individuals with chronic stroke. Both left and right step length targets were adjusted to achieve the correct step length asymmetry while keeping the summation of left and right step lengths constant. (C) An example of a step that hit the target. As the right leg moved forward, the gray bar grew proportionally. The participant’s heel strike occurred when the gray bar reached the step length target, and the target exploded to indicate correct step length. The yellow bar for the right leg changed position to show where the step length occurred for the next right step length. (D) An example of a step that missed the target. As the participant moved the left leg forward, the left gray bar grew proportionally. The participant’s heel strike was before the target, and the target turned red. The yellow bar was adjusted to show the left step length, indicating to the participant to take a longer left step.

During the last three minutes, the number of targets hit was displayed to further motivate participants. Different step length asymmetries were achieved by adjusting the step length targets while keeping the summation of paretic and nonparetic step lengths (or left and right step lengths for the unimpaired participants) constant to enforce a fixed stride length across conditions.

### Demographics of participants with chronic stroke

Twenty-eight individuals with chronic stroke (>6 months) were enrolled in this study, of which ten met the inclusion criteria (Table 1). To participate in this study, individuals had to: be capable of walking unassisted, aside from the use of an ankle foot orthosis; have sustained only one previous stroke; and have no other neurological disorders. We were interested in identifying the relationship between step length asymmetry and metabolic cost in individuals with chronic stroke who self-selected a clinically relevant step length asymmetry when they had the capacity to voluntarily alter asymmetry. Therefore, participants were excluded from the study if: they did not have a clinically meaningful step length asymmetry, defined as at least 4%, which is greater than the average step length asymmetry of unimpaired individuals(2); they were unable to alter step length asymmetry with biofeedback; they felt uncomfortable walking on the treadmill; or they were unable to walk without holding the treadmill handrail.

**Table 1.** Demographics for participants with chronic stroke.

| Participant | Sex | Age | Mass (kg) | Height (m) | Months since stroke | Lesion Type | Affected Side | Dominant Leg | LEFM | Mini-mental (Max 30) | TUG (s) | Maximum Overground Speed (m/s) | Selected Speed (m/s) | Criterion for Speed Selection | Self-selected Asymmetry (%±STDEV) | AFO |
| --- | --- | --- | --- | --- | --- | --- | --- | --- | --- | --- | --- | --- | --- | --- | --- | --- |
| P09 | Male | 61 | 90 | 1.76 | 17 | Left MCA | R | R | 32 | 14 | 11.3 | 1.28 | 0.96 | RER | 8.1 ± 1.1 | N |
| P12 | Female | 67 | 54 | 1.61 | 62 | Left MCA, frontal, parietal lobe and Basal Ganglia | R | R | 21 | 30 | 10.0 | 0.86 | 0.35 | RER | 18.4 ± 5.5 | N |
| P14 | Male | 52 | 98 | 1.75 | 254 | Frontotemporal parietal | R | R | 31 | 25 | 11.7 | 0.83 | 0.37 | Pain | -11.6 ± 3.7 | N |
| P16 | Male | 53 | 100 | 1.74 | 209 | Left Thalamus AVM | R | R | 17 | 24 | 12.4 | 0.84 | 0.41 | RER | 9.8* | Y |
| P19 | Male | 47 | 121 | 2.03 | 370 | Left frontal temporal | R | R | 32 | 23 | 7.2 | 1.74 | 1.2 | Handrail holding | -8.3 ± 1.1 | N |
| P22 | Male | 51 | 74.5 | 176.5 | 61 | Discarotid artery, right MCA/ACA | L | R | 18 | 21 | 22.9 | 0.54 | 0.22 | RER | -46.5 ± 3.5 | Y |
| P24 | Male | 77 | 93 | 1.79 | 35 | Left periventricular, temporal and Basal Ganglia | R | R | 33 | 26 | 8.6 | 1.24 | 0.51** | RER | 5.3 ± 0.8 | N |
| P26 | Female | 62 | 66 | 1.61 | 467 | Left frontal | R | L | 26 | 24 | 9.3 | 1.01 | 0.76 | RER = 0.8 | -7.8 ± 0.4 | N |
| P27 | Female | 56 | 84 | 1.63 | 76 | Left N/A | R | R | 18 | 25 | 22.2 | 0.39 | 0.13 | RER | -43.4 ± 7.2 | Y |
| P28 | Female | 55 | 72 | 1.655 | 86 | Left MCA | R | R | 26 | 28 | 12.2 | 0.83 | 0.45 | RER = 0.8 | 14.2 ± 2.4 | N |

\* Participant only attended one testing session
\*\* Experimenter error in speed; calculated speed was 0.64 m/s

### Demographics of unimpaired participants

Ten unimpaired individuals participated in the study as the control group (Table 2). To be included in the study, participants had to have no history of neurological disorders and be comfortable walking on a treadmill without holding the handrails.

**Table 2.** Demographics for unimpaired participants.

| Participant | Sex | Age | Mass (kg) | Height (m) | Dominant Leg | Self-selected Asymmetry (%) | Repeat Condition |
| --- | --- | --- | --- | --- | --- | --- | --- |
| U1 | Male | 25 | 93 | 1.82 | R | 1.1 | +12% |
| U2 | Male | 38 | 83 | 1.85 | R | 0.5 | -12% |
| U3 | Female | 25 | 59 | 1.70 | R | 0.4 | 0% |
| U4 | Female | 31 | 66 | 1.63 | R | 0.8 | -6% |
| U5 | Female | 23 | 61 | 1.71 | R | -0.5 | 0% |
| U6 | Female | 24 | 48 | 1.64 | L | 1.6 | -12% |
| U7 | Female | 23 | 53 | 1.65 | R | 0.9 | +6% |
| U8 | Male | 24 | 91 | 1.88 | R | -0.4 | -6% |
| U9 | Male | 24 | 72 | 1.71 | R | -0.3 | No BF |
| U10 | Male | 23 | 83 | 1.87 | L | -0.0 | +6% |

### Testing Paradigm for participants with chronic stroke

Individuals with chronic stroke participated in four experimental sessions over four days. The first session included treadmill and biofeedback familiarization, exclusion criteria implementation, and clinical testing (Figure 2 S1). Sessions two through four consisted of refamiliarization with the biofeedback system and data collection (Figure 2 S2-S4). Individuals with chronic stroke experienced three biofeedback conditions which targeted self-selected asymmetry, zero asymmetry, and exaggerated asymmetry, defined as twice their self-selected asymmetry. Rest breaks were given between every walking bout until blood pressure and heart rate returned to baseline measurements, typically about five minutes.

**Figure 2.**
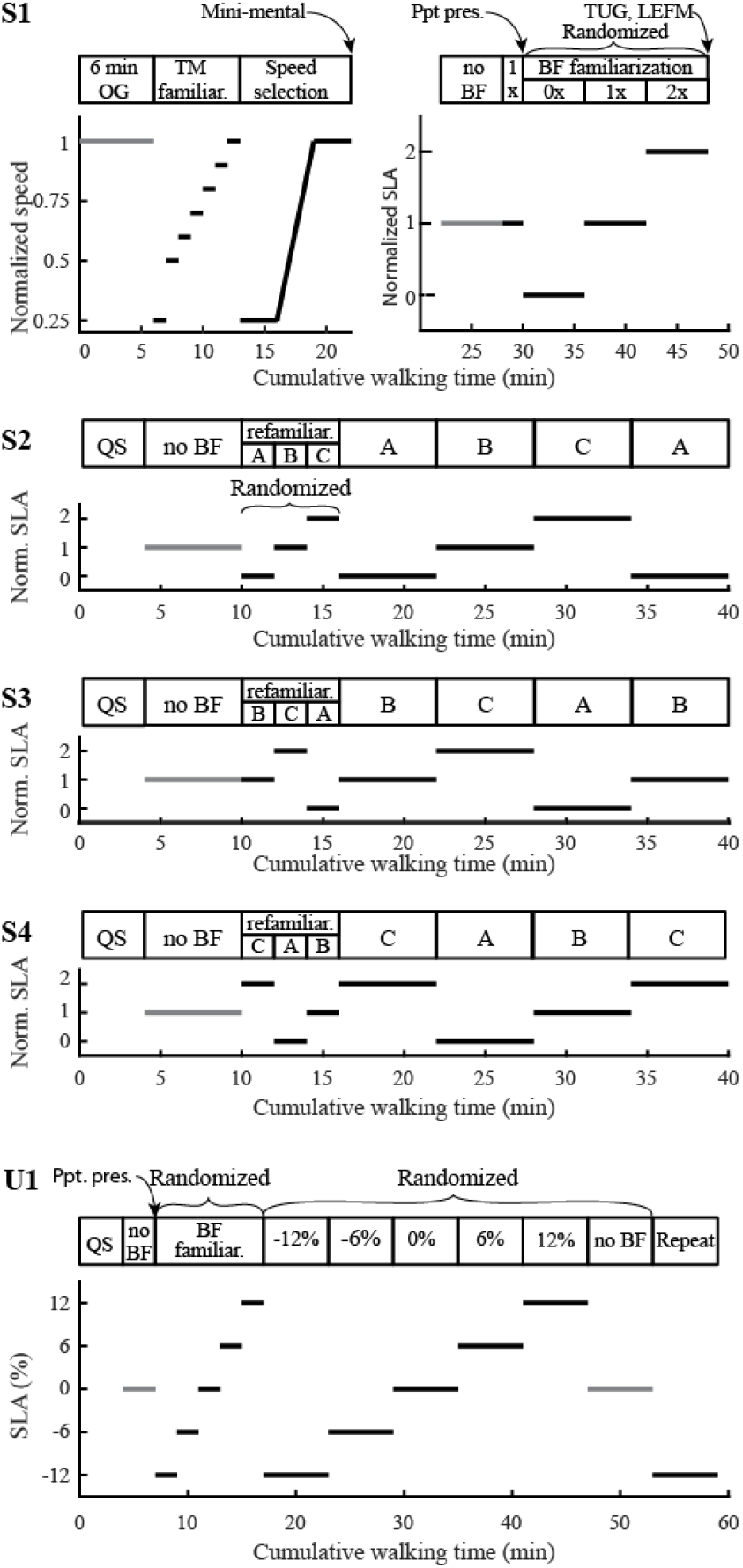
Testing paradigm. S1-S4 depict the sessions for participants with chronic stroke. U1 depicts the session for unimpaired participants. (S1) The first session for individuals with chronic stroke included treadmill familiarization, speed selection based on biological measurements, biofeedback training, and clinical testing. (S2) Resting metabolic rate was measured in quiet standing. Participants walked with no biofeedback initially to find left and right step lengths. The order of conditions was randomized, and participants were refamiliarized in the experimental order. Participants then walked in each condition for six minutes, and metabolic rate was measured. The first condition was repeated to reduce the effects of continued learning in the first condition. (S3 and S4) The same experimental protocol was conducted as S2, except the conditions were reordered. (U1) Experimental session for unimpaired participants. Resting metabolic rate was measured in quiet standing. Initial no biofeedback condition was used to find left and right step lengths for biofeedback. Participants were then trained in each condition. The order of conditions was randomized, and the first condition was repeated.

#### Familiarization and speed selection (Session 1)

The first session involved selecting an appropriate speed for each participant and familiarizing participants with treadmill walking and the biofeedback system. Participants first completed a six-minute walk test to determine their maximum average over-ground walking speed(38,39). Participants were then familiarized with treadmill walking. During treadmill familiarization (TM familiar. in Figure 2 S1), participants first walked at 25% of their maximum over-ground speed. The speed was then increased to 50% of their over-ground speed, and then gradually increased to 100% in 10% increments, with each speed held for about 1 minute or until the participant was comfortable with increasing the speed, indicated through verbal consent. The speed selection protocol (described in the section “Speed selection for participants with chronic stroke”) was then conducted to identify a speed that was comfortable and sustainable, based on biological measurements, and at which participants could still alter step length asymmetry.

Self-selected asymmetry was then measured at the selected speed. Participants walked on the treadmill at the selected speed for six minutes. To reduce the effects of outlier steps, the median asymmetry of the last three minutes of walking was used as the self-selected step length asymmetry. Participants who did not exhibit a self-selected step length asymmetry of at least 4% at the selected speed were excluded from the rest of the study.

Asymmetric participants were then trained on the biofeedback system. A slide presentation explaining the task was given to participants. To translate the instructions to the actual task, participants were first exposed to two minutes of walking while targeting their self-selected asymmetry using the biofeedback system described previously. Participants then trained for six minutes in each biofeedback condition (zero, self-selected, and exaggerated) presented in a randomized order. Participants were encouraged to ask questions throughout training. Verbal reinforcement and coaching were provided to participants to help improve accuracy and motivation. No metabolic data were collected during training to allow for easier communication. If participants were unable to alter their step length asymmetry with biofeedback, they were excluded from the rest of the experiment.

Clinical tests were also applied during the first session to determine the level of neurological impairment. A mini-mental cognitive test was conducted following speed selection. A timed up and go test (TUG) and a lower extremity Fugl-Meyer (LEFM) assessment were conducted at the end of the session.

#### Refamiliarization and data collection trials (Sessions 2-4)

Once individuals were familiarized with the task in the first session, metabolic rate and kinematic data were collected over three sessions. Asymmetry conditions were presented in pseudorandom order (Figure 2 S2-S4) to reduce potential ordering effects on metabolic cost. All walking bouts were on the treadmill at the speed selected during the first session. Each session began with a four-minute quiet standing trial to measure resting metabolic rate. Participants then walked for six minutes without biofeedback while their self-selected asymmetry and baseline metabolic rate for the session were measured. Participants were subsequently given two minutes of refamiliarization in each of the three biofeedback conditions in the experimental order of the session. Following refamiliarization, participants walked in each asymmetry condition, with biofeedback, for six minutes. Metabolic rate, kinematic, and kinetic data were measured simultaneously. The order of the three biofeedback conditions was randomized during session two, and the first condition was repeated to assess whether trial order had an effect during the course of the session. Only data from the second exposure were included in the analysis. The randomized order of conditions during session two was labeled as A-B-C-A. During sessions three and four, the order of the conditions was reordered to B-C-A-B and C-A-B-C, respectively, so that all conditions would be presented first to ensure equal exposure to all conditions and reduce potential ordering effects on metabolic cost.

### Testing paradigm for unimpaired participants

Unimpaired individuals participated in one session which included biofeedback familiarization and data collection (Figure 2 U1). One session was used because unimpaired individuals exhibited consistent trends over multiple days during pilot testing. Unimpaired participants completed symmetric and exaggerated walking conditions using the same biofeedback system as the participants with chronic stroke. Exaggerated asymmetries were based on the measured step length asymmetry in participants with chronic stroke. Short breaks were given between each condition to reset the equipment.

#### Familiarization and speed selection

Unimpaired individuals were familiarized with the biofeedback system but did not require treadmill familiarization given their experience walking on a treadmill. All participants walked at 1.25 m/s, a comfortable speed for unimpaired individuals walking on a treadmill(40). The experiment began with a four-minute quiet standing trial to measure resting metabolic rate. Unimpaired participants then walked for three minutes without biofeedback to calculate self-selected step lengths for the biofeedback system. Self-selected step lengths for the right and left legs were taken as the median values from the last minute of the trial.

Unimpaired participants were then familiarized with the biofeedback system. Participants were first given a slide presentation. Participants were then exposed to each of the five biofeedback conditions for two minutes in random order. They were encouraged to ask questions during the familiarization period.

Different target asymmetries were used in the unimpaired group because self-selected asymmetry was typically close to 0%. Target asymmetries were based on the step length asymmetries measured among the participants with chronic stroke. To select target asymmetries, we first found the absolute change in measured step length asymmetry between the self-selected with biofeedback condition and both the zero and exaggerated conditions for participants with chronic stroke. The smaller of the two changes in asymmetry for each experimental session was used for analysis. The median of this change across all participants and sessions was about 6%. We therefore selected -12%, -6%, 0, +6%, +12% asymmetry conditions for the unimpaired participants.

#### Data collection trials

Unimpaired participants walked for six minutes in each of the five biofeedback conditions and in one no biofeedback condition. Conditions were presented in random order. The first condition they were exposed to was repeated at the end of the session. Only data from the second exposure were included in the analysis. Metabolic rate, kinematic, and kinetic data were measured during each trial.

### Speed selection for participants with chronic stroke

We developed and applied an elaborate protocol to identify the speed at which each participant with chronic stroke could walk for six minutes while modulating step length asymmetry without exceeding limits on heart rate or respiratory exchange ratio (RER). Participants walked on a treadmill for a maximum of nine minutes during the speed selection protocol. Metabolic rate, heart rate, and respiratory exchange ratio (the ratio of carbon dioxide production to oxygen consumption) were collected. We then performed calculations to determine the speeds at which heart rate exceeded a safe value or respiratory exchange ratio indicated the potential for anaerobic energy consumption (Figure 3).

**Figure 3.**
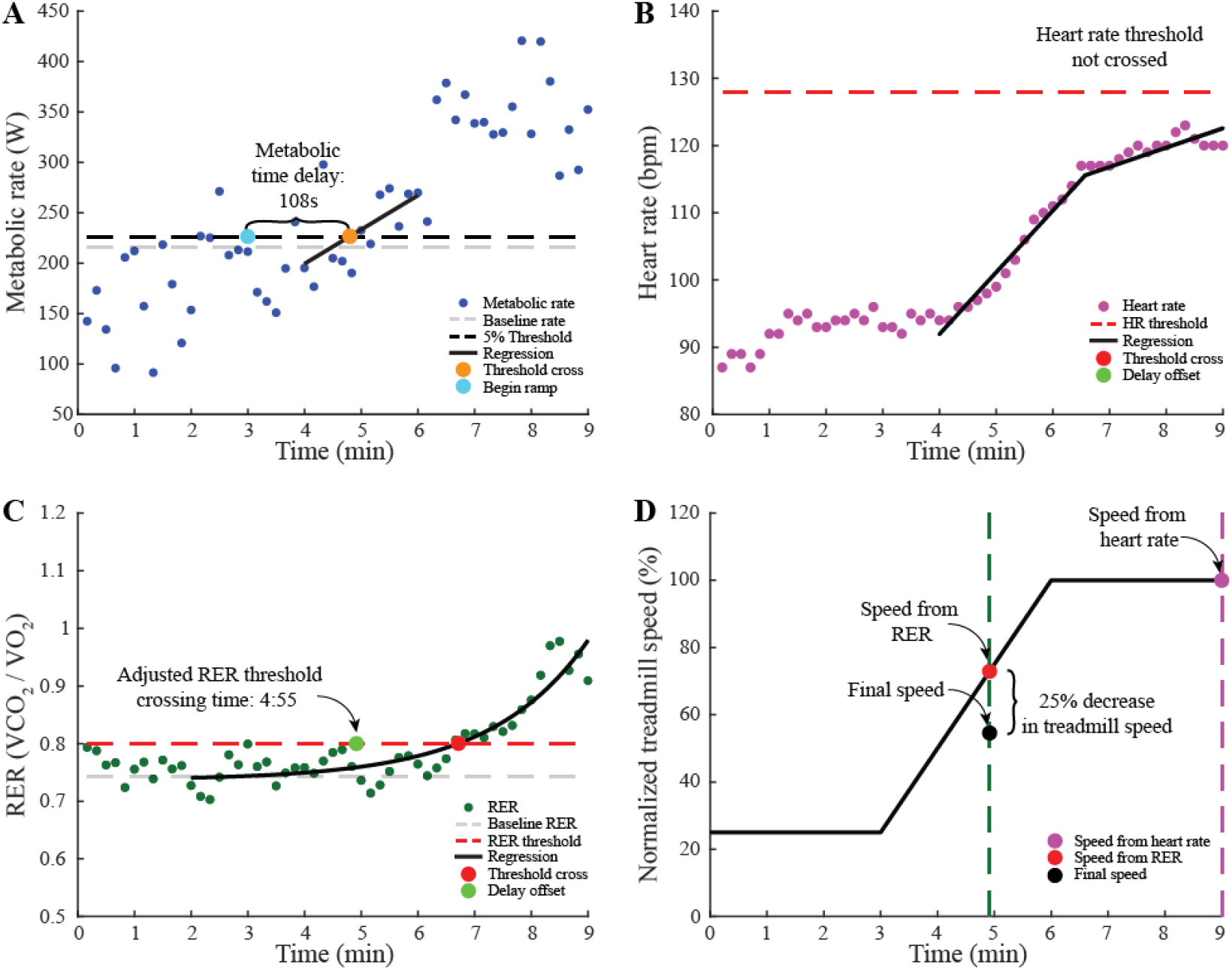
Example speed selection protocol for participants with chronic stroke. (A) Metabolic rate used to find time delay in body’s response time. The baseline metabolic rate was the average rate between minutes two and three. Data were fit with linear regression between minutes four to six. The time the regression crossed the 5% increase in metabolic rate was found. The time delay was the difference between the threshold crossing and the onset of the treadmill speed increase. (B) Linear regressions on heart rate data to find time when heart rate reaches the 78% maximum heart rate threshold. The time delay was subtracted from the time the heart rate regression crossed the threshold. The heart rate did not cross the threshold in this participant. (C) Exponential model fit between minutes two to nine on respiratory exchange ratio. The threshold for respiratory exchange ratio was the larger of 0.8 respiratory exchange ratio or a 5% increase from baseline respiratory exchange ratio. The time delay was subtracted from the time the exponential fit crossed the threshold. (D) Normalized treadmill speed throughout the protocol to map threshold times to speed. 75% of the lower speed between respiratory exchange ratio and heart rate was the speed selected for participants with chronic stroke.

In the first three minutes, participants walked at 25% of the over-ground speed measured in the six-minute walk test to collect their baseline biological measurements. Treadmill speed was then linearly increased to 100% of their over-ground speed during the next three minutes. For the final three minutes, speed was held constant at the participant’s over-ground speed to test whether the participant could sustain that speed. The speed selection was stopped if: the participant’s heart rate exceeded 90% of their maximum heart rate; their respiratory exchange ratio reached 1.1 indicating anaerobic respiration; they could not walk without holding the handrail; or they requested to stop because of pain.

We then calculated the delay in metabolic response to the increasing treadmill speed (Figure 3A). The baseline metabolic cost of walking was computed as the average cost during minutes two and three. As treadmill speed linearly increased, metabolic rate also increased, but with a time delay(28). This time delay was approximated by fitting a linear regression to the metabolic cost during minutes four to six. The delay was quantified as the time difference between the onset of the speed increase at minute three and the time when the linear regression reached a 5% increase from the average baseline metabolic rate.

The maximum sustainable speed was determined using thresholds on heart rate and respiratory exchange ratio. Linear regressions were fit to the heart rate data between minutes four to six and seven to nine because of the different speed profiles (Figure 3B). Based on pilot testing, we set the threshold for heart rate at 78% of the participants’ maximum heart rate, defined as 220 bpm minus their age in years. An exponential curve was fit to the respiratory exchange ratio data between minutes two and the end of the trial (Figure 3C). The baseline respiratory exchange ratio was calculated as the average of the exponential curve between minutes two and three. The threshold for respiratory exchange ratio was the larger of either a 5% increase from the baseline respiratory exchange ratio or a respiratory exchange ratio of 0.8. The times when the fits of heart rate and respiratory exchange ratio reached the thresholds were calculated, and the metabolic time delay was subtracted. Treadmill speed at the earlier of the two times was determined (Figure 3D). Based on pilot tests, we found than an additional 25% decrease in speed was required to allow for increased exertion when modulating step length asymmetry.

### Measurements collected

#### Kinematic and kinetic data

Ankle locations were collected by tracking reflective markers attached to the lateral malleoli with a passive motion analysis system sampling at 100 Hz (Vicon Motion Systems, Oxford, UK). Vertical ground reaction forces measured by the instrumented treadmill at 1000 Hz (Bertec Corporation, Columbus, OH) were used to detect heel strike (Fz > 10N) and toe-off (Fz < 10N).

#### Metabolic rate and heart rate

Metabolic rate was indirectly calculated by measuring the rate of oxygen consumed and rate of carbon dioxide produced using a mobile respirometry system (Oxycon Mobile; CareFusion, San Diego, CA or Cosmed K4b2, Rome, Italy) and applying a standard equation(41). Respiratory exchange ratio was also calculated using these data. Heart rate was measured using a transmitting electrocardiogram (T31 Coded Transmitter; Polar, Kempele, Finland).

### Data processing and analysis

#### Step length asymmetry

Step length asymmetry was calculated using the location of the ankles at each heel strike. Step length was defined as the distance between the leading- and trailing-leg ankle markers at heel strike. Paretic and nonparetic step lengths were defined when the respective leg was leading at heel strike.

Step length asymmetry was calculated as the normalized difference between the step lengths of the paretic and nonparetic limb(42,43):

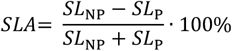

where SLA is step length asymmetry, SL_p_ is the step length of the paretic leg, and SL_NP_ is the step length of the non-paretic leg. For unimpaired controls, left and right step lengths were used instead of paretic and nonparetic step lengths. Step length asymmetry was measured by averaging the step length asymmetries over the last three minutes of each walking bout.

#### Metabolic cost

Volumetric flow rates were measured breath-by-breath and averaged over ten second intervals. We used the average metabolic rate of the last three minutes of each condition. Net metabolic rate was calculated by subtracting the average metabolic rate during quiet standing from the average metabolic rate during each walking condition.

#### Normalization of data and statistical analysis

For the population with chronic stroke, step length asymmetry was normalized to self-selected asymmetry without biofeedback. Step length asymmetry was not normalized for the unimpaired controls because their self-selected asymmetry was close to 0%. Metabolic rate was normalized to body mass for each participant. Percent change in metabolic rate was calculated by comparing the average metabolic rate measured during each biofeedback condition to the average metabolic rate of the self-selected asymmetry without biofeedback condition. Each session was normalized separately. In all statistical analyses, only the biofeedback conditions were compared.

Statistical comparisons were made for step length asymmetry and metabolic rate across biofeedback conditions. Average percent change in metabolic rate and measured step length asymmetry for each condition were tested for normality using the Kolmogorov-Smirnov test. To test whether measured step length asymmetry and metabolic rate differed across biofeedback conditions, we performed three-way ANOVA (random effects: participant and session; fixed effect: biofeedback condition, treated as categorical). If significance was found, paired t-tests were performed to compare biofeedback conditions. We also tested whether metabolic rate was correlated with measured step length asymmetry according to a quadratic model, using regression analysis. Significance for comparisons was set at α = 0.05.

## Results

Participants in both groups significantly altered their step length asymmetry with biofeedback. Metabolic rate was not different across conditions in individuals with chronic stroke. Metabolic rate significantly changed across step length asymmetry conditions in unimpaired controls.

### Step length asymmetry modulation

#### Participants with chronic stroke

Significant changes in step length asymmetry were observed across biofeedback conditions for participants with chronic stroke (ANOVA; F_2,70_ = 173, p = 8e-28; Figure 4A). Across the group, individuals with chronic stroke significantly altered their step length asymmetry with biofeedback from an average normalized asymmetry of 1.0 (average absolute asymmetry 18% ± 17%) in the self-selected condition to 0.4 (7.3% ± 9.7%) in the symmetry condition and 1.5 (27% ± 22%) in the exaggerated condition (paired t-tests, n = 10, p = 4e-14 and p = 6e-11, respectively). Large standard deviations in absolute asymmetry were due primarily to inter-subject variability.

**Figure 4.**
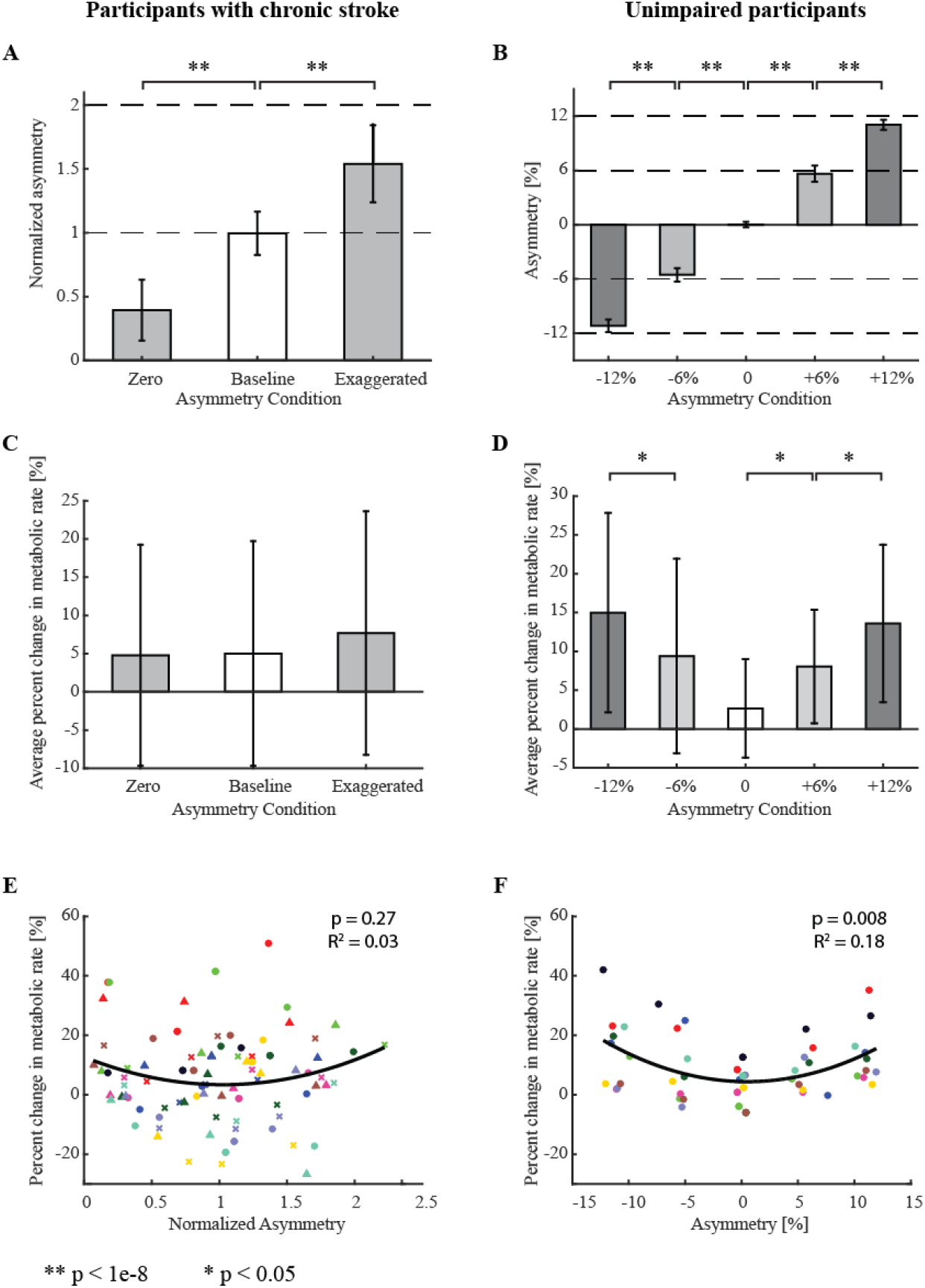
Step length asymmetry modulation and metabolic rate results. Results for participants with chronic stroke are on the left and those for unimpaired participants are on the right. (A) Average measured step length asymmetry across participants with chronic stroke. Asymmetry was normalized to self-selected without biofeedback. (B) Average measured step length asymmetry for unimpaired participants. Participants in both groups significantly altered step length asymmetry from self-selected as a result of study design. (C) Average percent change in metabolic rate compared to the self-selected without biofeedback condition. Metabolic rate was unchanged across conditions among participants with chronic stroke. (D) Metabolic rate increased with increases in absolute step length asymmetry among unimpaired participants. (E) Metabolic rate versus measured step length asymmetry for all trials among individuals with chronic stroke. Very little of the change in metabolic rate was explained by step length asymmetry. Different colors represent different participants, and different shapes represent different experimental sessions. (F) A significant trend between metabolic rate and measured step length asymmetry was found for unimpaired participants. Different colors represent different participants.

#### Unimpaired participants

Unimpaired participants significantly altered step length asymmetry across biofeedback conditions (ANOVA; F_4,36_ = 6.19, p = 7e-4; Figure 4B). Step length asymmetry differed between the -12% and -6% conditions (absolute difference of 5.6%, paired t-test, n = 10, p = 5e-10), -6% and 0% conditions (5.4%, p = 4e-9), 0% and 6% conditions (5.7%, p = 2e-8), and 6% and 12% conditions (5.5%, p = 5e-9).

### Metabolic rate comparisons

#### Participants with chronic stroke

Percent change in metabolic rate was not found to be different across targeted biofeedback conditions (ANOVA, F_2,70_ = 0.39, p = 0.68; Figure 4C). The quadratic regression between percent change in metabolic rate and measured asymmetry was not significant (p = 0.27, R^2^ = 0.03; Figure 4E).

#### Unimpaired participants

Percent change in metabolic rate was different across targeted biofeedback conditions (ANOVA, F_4,36_ = 6.25, p = 6e-4; Figure 4D). Post hoc analysis revealed that higher asymmetry resulted in higher metabolic rate comparing the -12% and -6% conditions (5.6% greater, p = 0.028), the -6% and 0% conditions (6.1% greater, p = 0.078), 6% and 0% (6.1% greater, p = 0.012), and 12% and 6% (5.6% greater, p = 0.04), The quadratic regression between percent change in metabolic rate and measured asymmetry was significant (p=0.008, R^2^=0.18; Figure 4F).

## Discussion

The purpose of this study was to test whether minimizing metabolic rate could explain self-selected step length asymmetry in individuals with chronic stroke. We conducted a study in which we asked participants to alter their step length asymmetry while measuring their metabolic cost. We found that minimizing metabolic cost does not seem to explain self-selected step length asymmetry in participants with chronic stroke; no differences in metabolic rate were seen across asymmetry levels. To test whether the lack of relationship between step length asymmetry and metabolic rate was related to stroke, we performed a similar experiment on unimpaired individuals. Unimpaired participants self-selected a low step length asymmetry which corresponded to their energy minimum. These findings suggest that, after a stroke, factors other than metabolic rate minimization drive self-selected step length asymmetry.

### Metabolic rate

Previous studies have shown that changing gait parameters affects metabolic rate in unimpaired participants. People will usually self-select a gait near their energy minimum, and small changes in their gait typically lead to increases in metabolic cost(19). In this study, unimpaired participants exhibited the expected change in metabolic rate with changing step length asymmetries, where changes in asymmetry led to increases in metabolic rate. The participants with chronic stroke included in this study were able to change their step length asymmetry but exhibited a qualitatively different metabolic response. As a group, participants with chronic stroke had similar metabolic cost across step length asymmetry conditions, consistent with previous studies(20,29).

Some gait parameters, such as walking speed(44), have energetic cost of transport landscapes with a shallow slope near the energy minimum. In part because the metabolic penalty for walking at speeds near the minimum is small, unimpaired individuals walk with a large variation in self-selected speed(45). While we did not find a significant relationship between metabolic rate and asymmetry in individuals with chronic stroke, a shallow energy landscape might exist. This would be difficult to identify given the high variability in kinematics(46) and energy cost exhibited by individual participants walking under the same asymmetry conditions. If a shallow landscape does exist, we do not expect metabolic cost to be the main factor driving step length asymmetry in participants with chronic stroke. If participants with chronic stroke can walk with a wide range of asymmetries without substantially affecting metabolic rate, other optimization goals could dominate, such as increasing stability(47), avoiding fatigue(48,49), or reducing single-support time on their paretic limb(3), resulting in the observed self-selected step length asymmetry.

Muscle fatigue avoidance has been shown in simulation to explain some gait patterns in older adults(49), and it might explain self-selected step length asymmetry in individuals with chronic stroke. Previous studies have shown that to reduce step length asymmetry, individuals with chronic stroke increase paretic ankle plantarflexor activation(50,51). Paretic plantarflexors might fatigue faster than the nonparetic muscles, so participants could be walking asymmetrically to reduce muscle fatigue.

Other forms of asymmetry contribute to step length asymmetry, such as asymmetries in paretic propulsion(25), step position(42), or step timing(21,42). Individuals could have altered step length asymmetry by changing any of these factors, some of which could affect metabolic cost(21). Future studies providing specific feedback to isolate other asymmetries are necessary to examine their correlation with metabolic cost.

The metabolic penalty for deviating from self-selected step length asymmetry is larger for unimpaired participants than for participants with chronic stroke. Differences in energy landscape could be due to the differences in speed between these populations. If participants with chronic stroke walked faster, a steeper energy landscape might be found. However, none of the participants with chronic stroke, including two that walked at a speed similar to the walking speed of the unimpaired participants, exhibited a consistent energy landscape.

### Step-length asymmetry modulation

While both populations in this study altered their self-selected step length asymmetry, participants with chronic stroke did so to a lesser extent than we requested compared to unimpaired participants. Participants with chronic stroke walked with step length asymmetries farther from the targeted values than the unimpaired participants, suggesting that individuals with chronic stroke had more trouble modulating step length asymmetry. This could be explained by the fact that individuals with chronic stroke have less complex coordination patterns than their unimpaired counterparts(52). The decrease in complexity could reduce the number of coordination strategies available to alter step length asymmetries as more neural signals have become coupled, leading to the observed difficulties.

Participants with chronic stroke maintained consistent self-selected step length asymmetry across sessions. Since consistent self-selected step length asymmetry in participants with chronic stroke cannot be explained by energy cost minimization, other factors such as stability(47) muscle weakness(53), or fatigue avoidance(48,49) must be responsible. For example, individuals with chronic stroke often experience diminished control(52), spasticity(23), or muscle weakness(53) of the paretic limb which could cause them to rely less on their paretic leg, leading to the observed consistent self-selected asymmetry. Additional studies should be conducted to identify which of these factors are most important to self-selected step length asymmetry following stroke.

Unimpaired participants walked with a step length asymmetry closer to their self-selected asymmetry in all biofeedback conditions, also indicating a step length asymmetry preference. In this case, trying to walk closer to their self-selected asymmetry in the biofeedback conditions can be explained through energy minimization. The more asymmetric unimpaired participants’ step lengths were, the more energy they used. They appeared to have tried to take advantage of the acceptable error allowed in the biofeedback task while still hitting the targets to decrease the effort needed to walk in each asymmetry condition.

### Limitations

A limitation of this study is that the unimpaired controls were not age, gender, or speed matched with their post-stroke counterparts. Therefore, we could not isolate the effects that stroke had on changes in metabolic rate and step length asymmetry. However, we wanted to understand if unimpaired adults self-select a step length asymmetry that minimizes their energy cost at a speed they might self-select for themselves to better understand asymmetry in unimpaired walking. The calculated speed for participants post-stroke was slower than their maximum over-ground speed. At faster speeds, asymmetry could have had a larger effect on metabolic rate. However, many participants with chronic stroke were unable to sustain altered step length asymmetry at faster speeds.

The biofeedback task might have been mentally challenging for individuals with chronic stroke, leading to mental fatigue or loss in motivation. We wanted participants to voluntarily alter their step length asymmetry rather than mechanically manipulate participants’ asymmetry using the split-belt treadmill, since voluntary modulation provided a better test of our hypothesis. When we noticed participants were frequently missing targets, we provided verbal encouragement to help maintain motivation. The challenge of voluntarily modulating step lengths could have increased variability in step length asymmetry and metabolic rate, thus reducing statistical power. Because we only altered step length asymmetry in this study, we were unable to determine if a relationship exists between metabolic energy consumption and other types of asymmetry, such as temporal, kinetic, or kinematic.

### Conclusions

We provided biofeedback to participants to encourage them to voluntarily alter step length asymmetry, and we measured their metabolic rate during each condition. We confirmed that unimpaired individuals self-select a near symmetric step length asymmetry that minimizes metabolic cost. When unimpaired participants walked more asymmetrically, their energy consumption increased. This is consistent with findings for many other parameters in unimpaired gait. Minimizing energy cost seems to influence walking behavior in unimpaired individuals.

In this study, participants with chronic stroke were able to voluntarily alter their step length asymmetry. We found that individuals with chronic stroke have a strong preference for their self-selected step length asymmetry, but we could not explain this preference with energy cost minimization. Additional insights into how participants altered their step length asymmetry could be gained from looking at changes in joint kinematics and muscle activity. To better understand why individuals with chronic stroke walk with their self-selected step length asymmetry, future studies should be conducted to examine other potential influences, such as balance enhancement or fatigue avoidance.

While minimizing energy use did not explain self-selected step length asymmetry in individuals with chronic stroke, altering metabolic energy consumption could still be a useful tool for rehabilitation. Our results suggest that because metabolic cost does not increase with altered asymmetry, metabolic energy consumption probably will not be a barrier to therapies involving practicing improved step length symmetry post-stroke. In fact, manipulating the energy landscape(19) to encourage symmetric step length asymmetries could be more easily accomplished. Rehabilitation methods that use changes in metabolic energy consumption to either incentivize a desirable step length asymmetry or penalize an undesirable step length asymmetry might encourage individuals with chronic stroke to self-select a more symmetric gait and improve long-term rehabilitation outcomes.

## Data Availability

Data will be available upon request to the corresponding author.

## Abbreviations

LEFM: Lower extremity Fugl-Meyer
TUG: Timed up and go
RER: Respiratory exchange ratio
SLA: Step length asymmetry
SL: Step length
NP: Nonparetic
P: Paretic

## Declarations

### Ethics approval, consent to participate, and consent for publication

All participants provided written informed consent for participation and publication before participating. The experimental protocol was approved by the University of Pittsburgh Institutional Review Board.

### Availability of data and materials

The datasets from this study are available in the supplemental materials

### Authors’ contributions

S.H.C. and G.T.O. conceived and oversaw the study; T.M.N., Y.A., R.W.J., and D.K. conducted data collections; T.M.N. and Y.A. analyzed data; T.M.N. led manuscript preparation; all authors edited the manuscript and approved the final manuscript.

## Acknowledgements

The authors would like to thank William Anderton and Nicolas Velasquez for help with participant recruitment and data collection. We thank to Bailey Patterson and Carly Sombric for help with data collection and Kirby Witte for help with pilot testing and biofeedback development.

